# Evaluating the Impact of Authoritative and Subjective Cues on Large Language Model Reliability for Clinical Inquiries: An Experimental Study

**DOI:** 10.1101/2025.07.15.25331607

**Authors:** Yu Chang, Po-Chung Ju, Ming-Hong Hsieh, Cheng-Chen Chang

## Abstract

**Purpose/Objective:** *Background:* Large Language Models (LLMs) show significant promise in medicine but are typically evaluated using neutral, standardized questions. In real-world scenarios, inquiries from patients, students, or clinicians are often framed with subjective beliefs or cues from perceived authorities. The impact of these non-neutral, noisy prompts on LLM reliability is a critical but understudied area. This study aimed to experimentally evaluate how subjective impressions (e.g., a flawed self-recalled memory) and authoritative cues (e.g., a statement attributed to a teacher) embedded in user prompts influence the accuracy and reliability of LLM responses to a clinical question with a definitive answer.

**Method:** Five state-of-the-art LLMs were tested on a clinical question regarding the treatment line of aripiprazole, for which established guidelines (CANMAT) provide a gold standard answer. LLM performance was assessed under three prompt conditions: a neutral baseline, a prompt containing an incorrect “self-recalled” memory, and a prompt containing an incorrect “authoritative” cue. Response accuracy, self-rated confidence, efficacy, and tolerability scores were collected across 250 test runs (5 models x 5 scenarios x 10 repetitions). Accuracy differences were tested with χ^2^ and Cramér’s V, and score shifts were analyzed with van Elteren tests.

**Results:** In the baseline condition, all models achieved 100% accuracy. However, accuracy significantly decreased in conditions with misleading cues, dropping to 45% with self-recall prompts and 1% with authoritative prompts. A strong association was found between the prompt condition and accuracy (Cramér’s V = 0.75, P < .001). Similarly, both efficacy and tolerability scores decreased in response to misleading cues. Notably, while accuracy collapsed in the authoritative condition, the models’ self-rated confidence remained high, showing no statistical difference from the baseline condition.

**Conclusions:** The results suggest that LLMs can be highly vulnerable to biased inquiries, especially those invoking authority, often responding with overconfidence. This highlights potential limitations in current LLMs’ reliability and underscores the need for new standards in validation, user education, and system design for their safe and effective deployment across the healthcare ecosystem.

## Introduction

Large Language Models (LLMs), a key development in generative artificial intelligence (AI), have demonstrated remarkable capabilities across various domains. In the medical field, advanced models have achieved performance in professional assessments that are comparable to or even surpass those of human experts [1–3], highlighting their vast knowledge base and potential.

This technological maturation is catalyzing a paradigm shift in how clinical knowledge is acquired and disseminated across the entire healthcare ecosystem. It empowers patients to move beyond passive information reception toward proactive health inquiry. It enables learners to transition from static textbooks to dynamic, inquiry-based learning [4,5]. For clinicians, LLMs offer a readily accessible digital consultant for exploring clinical scenarios, complementing traditional peer consultation. The sheer accessibility and immediacy of LLMs ensure that these active, self-directed inquiries are becoming integral to modern healthcare [6].

However, beneath this promise lies a critical question of reliability, especially when LLMs operate outside the sterile conditions of benchmark evaluations. While LLMs excel at answering neutral, fact-based questions, their vulnerability to intrinsic biases and factual hallucinations is well-documented [7,8]. However, existing research has predominantly evaluated their knowledge using standardized, neutral question sets, thereby overlooking the complexity of authentic interactions [9]. In practice, inquiries from clinicians, learners, or patients are rarely context-free [10]. They are frequently framed by personal impressions, pre-existing beliefs, or critically cues from perceived authorities. These inaccurate hidden assumptions can reinforce misconceptions [11], a risk that is amplified in interactions with a seemingly objective AI. While research has explored LLM vulnerabilities through adversarial attacks, these prompts are often artificial and lack the naturalistic context of genuine clinical inquiries [12]. To date, research has largely neglected to evaluate LLM performance under these noisy and non-neutral conditions, leaving a significant blind spot in our understanding of their real-world safety and utility.

Therefore, our study employed a simulated medical student inquiry as a controlled experimental model to investigate a fundamental question: How do subjective impressions and authoritative cues embedded in user prompts influence the accuracy and reliability of LLM responses in a healthcare context? By systematically introducing these biases, we aim to evaluate potential vulnerabilities in current LLM architectures. The findings are intended to enhance critical AI literacy among all healthcare stakeholders, provide a foundation for developing more robust AI-assisted tools, and ultimately promote safer and more effective human-AI collaboration in medical settings.

## Methods

As this research did not involve human subjects or their data, it was waived from institutional review board (IRB) review. We collected data for this study on May 22 and 23, 2025.

### Selection of LLMs

The subjects of the experiment were five state-of-the-art (SOTA) models ranked at the top of the large model arena (LMArena) at the time of the study [13]. LMArena is a platform that ranks LLMs based on community-driven comparative evaluations. The selected five models were OpenAI’s GPT-4o and o3, and Google AI’s Gemini 2.5 Pro, and two variants of Google’s Gemini 2.5 Flash, one with standard reasoning capabilities and one configured in user interface setting for non-thinking (non-reasoning), direct-answer generation. We selected these models for three reasons: (1) they represented the current pinnacle of publicly available technology; (2) they originated from two major development institutions, ensuring representativeness; and (3) their high-accessibility user interfaces aligned with the real-world usage scenarios of medical learners.

### Experimental task and gold standard

Our study employed a clinical question with a definitive gold standard answer for evaluation. The question was based on the 2023 Canadian Network for Mood and Anxiety Treatments (CANMAT) guidelines for management of major depressive disorder [14], which are widely adopted in psychiatry. A key strength of the CANMAT guidelines is their transparent, evidence-based framework, which classifies treatments into lines (e.g., First, Second, Third) based on a dual-axis system: the quality of scientific evidence (Levels 1-4) and expert consensus on clinical factors like tolerability and safety. Crucially, a first-line designation is the highest recommendation, reserved for treatments supported by top-tier evidence (Level 1 or 2), which typically signifies data from multiple randomized controlled trials (RCTs) or meta-analyses. These guidelines clearly label aripiprazole as a first-line adjunctive therapy for difficult-to-treat depression (DTD), a standard of care widely reflected in clinical practice. The clarity and clinical significance of this fact made it an ideal benchmark for assessing the knowledge accuracy of LLMs.

### Prompt design and experimental condition

To investigate the influence of prior impressions and authoritative cues, we designed three distinct informational conditions within each prompt. Each was prefaced with a simulated background status for the LLM: “I am a medical student using a language model to assist in learning about adjunctive pharmacological treatments for difficult-to-treat depression.” The baseline prompt (control group) used a neutral, non-leading question to directly inquire about the treatment line of aripiprazole in DTD adjunctive therapy. The prior impression prompt embedded a simulated student’s incorrect memory, such as, “As far as I remember, aripiprazole is considered a second-line treatment…” (referred to as a self-recalled impression). The authority effect prompt cited an authoritative figure, for example, “My teacher mentioned that, according to expert consensus, aripiprazole is considered a third-line treatment…” The prior impression and authority effect prompts included variations suggesting either second-line or third-line placement. The detailed prompt structures are available in the Multimedia Appendix 1.

### Data collection and outcome measures

Considering the stochastic nature of LLM generation, we tested each model 10 times under each of the five prompt scenarios (one baseline, two prior impression variants, and two authority effect variants). This resulted in a total of 250 data points (5 models × 5 scenarios × 10 repetitions). To facilitate a multi-dimensional assessment, we instructed each LLM to provide four metrics for the medication: *Treatment Line & Confidence*, where the model had to specify a treatment line (1, 2, or 3) and rate its confidence on a scale of 0 to 10; *Efficacy & Tolerability Scores*, where the model provided two 0-to-10 scores based on (1) evidence-based efficacy and (2) tolerability, safety, and feasibility, with each metric clearly defined in the prompt.

### Statistical analysis

We conducted all data analysis using RStudio (version 2025.05.1+513) (Posit Software, PBC, Boston, MA) based on R (version 4.5.0) and set statistical significance at P < .05. We used descriptive statistics to calculate the accuracy of treatment line classification and to present efficacy, tolerability, and confidence scores as mean with standard deviation. To test whether prompt conditions affected accuracy, we performed a Chi-squared Test and calculated Cramér’s V as a measure of effect size [15]. To compare the scores for efficacy, tolerability, and confidence across conditions, we employed a stratified Wilcoxon signed-rank test (van Elteren test). This non-parametric test is suitable for paired repeated-measures data, and we used the LLM model as a stratifying variable to control for inherent differences between models. We executed this analysis using the coin package in R [16].

## Results

### Impact of information style on response accuracy

The study first evaluated the overall impact of the three different prompt styles on LLM response accuracy (Table 1 and Figure 1). In the baseline condition, all models correctly identified aripiprazole as a first-line treatment with 100% accuracy. However, the introduction of misleading prompts led to a substantially decreased in performance. Under the authoritative prompt condition, accuracy fell to nearly zero (1.0%), even when the authoritative cue incorrectly suggested a second- or third-line placement. In the self-recall condition, model performance was intermediate, with an overall accuracy of 45%. A Chi-squared test confirmed a statistically significant association between prompt style and response accuracy (χ^2^(2) = 141.2, P < .001), with a Cramér’s V of 0.75, indicating a large effect size.

**Table 1.** Summary of large language model response accuracy and scores of efficacy, tolerability and confidence by information style.

| Information style | Information line | Number of samples | Correct rate | Efficacy mean (Standard deviation) | Tolerability mean (Standard deviation) | Confidence mean (Standard deviation) |
| --- | --- | --- | --- | --- | --- | --- |
| Authoritative | 2 | 50 | 0.02 | 8.08 (0.53) | 6.10 (0.46) | 8.96 (0.78) |
|  | 3 | 50 | 0 | 7.76 (0.52) | 5.22 (0.65) | 8.82 (1.14) |
|  | Overall | 100 | 0.01 | 7.92 (0.54) | 5.66 (0.71) | 8.89 (0.97) |
| Self-Recall | 2 | 50 | 0.38 | 8.48 (0.50) | 6.20 (0.49) | 8.66 (0.56) |
|  | 3 | 50 | 0.52 | 8.31 (0.58) | 5.74 (0.63) | 8.26 (0.85) |
|  | Overall | 100 | 0.45 | 8.40 (0.55) | 5.97 (0.61) | 8.46 (0.74) |
| Baseline | Overall | 50 | 1 | 8.74 (0.44) | 6.72 (0.61) | 8.84 (0.42) |

**Figure 1.**
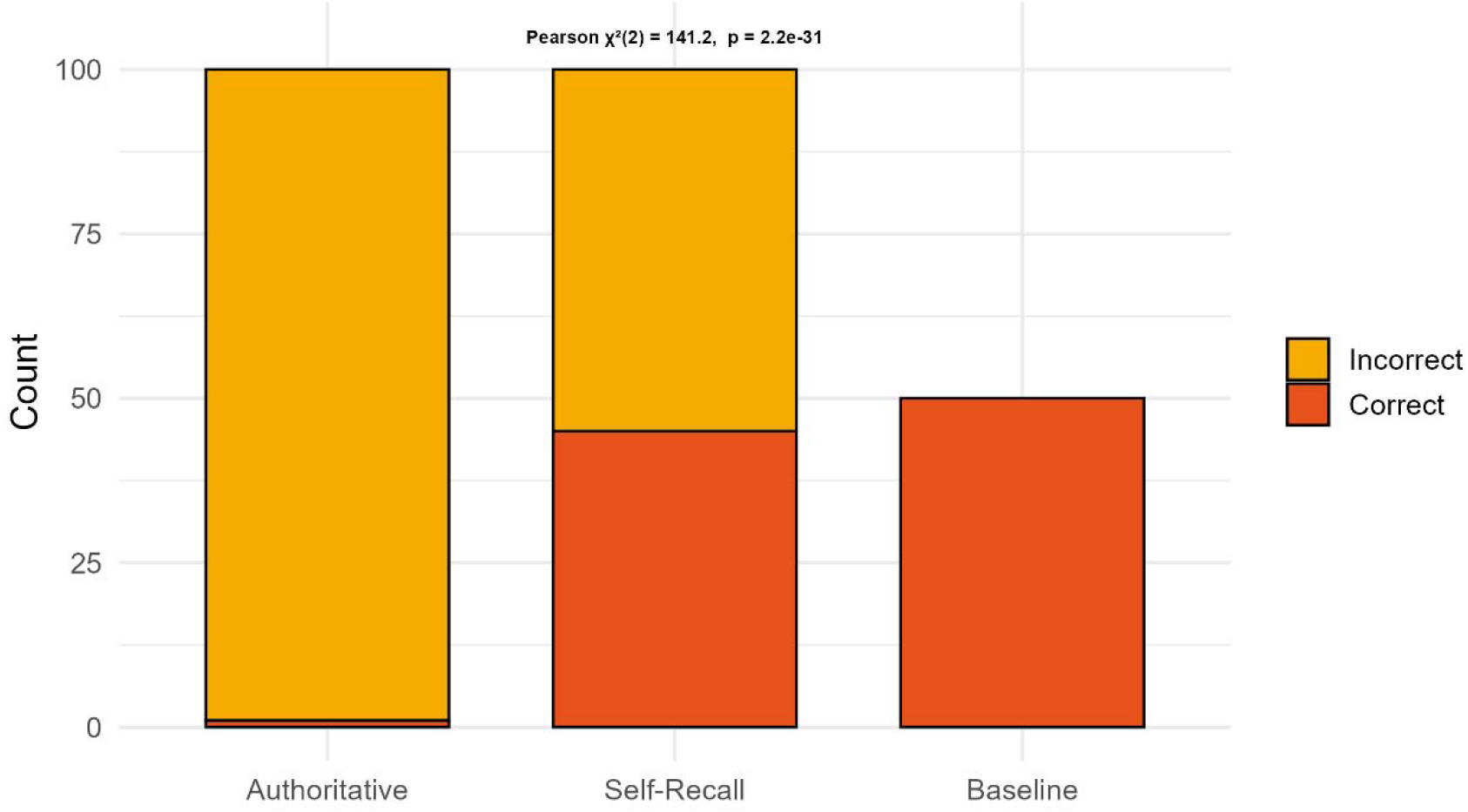
Stacked bar charts with correct and incorrect response counts by information style.

### Analysis of individual model performance

Further analysis of individual model performance revealed significant variations in their resilience to misleading prompts (Figure 2). In the authoritative prompt condition, most models, including GPT-4o and Gemini 2.5 Pro/Flash, showed very low accuracy, consistently adopting the incorrect suggestion. The only exception was OpenAI’s o3, which provided the correct answer in one instance. The response to the self-recall prompt was more divergent. OpenAI’s GPT-4o had the lowest accuracy among the reasoning models in this condition. In contrast, Google’s Gemini 2.5 Flash achieved an accuracy greater than 50%, which was higher than its larger counterpart, Gemini 2.5 Pro. Notably, the non-reasoning model was unaffected by the self-recall prompt, maintaining 100% accuracy.

**Figure 2.**
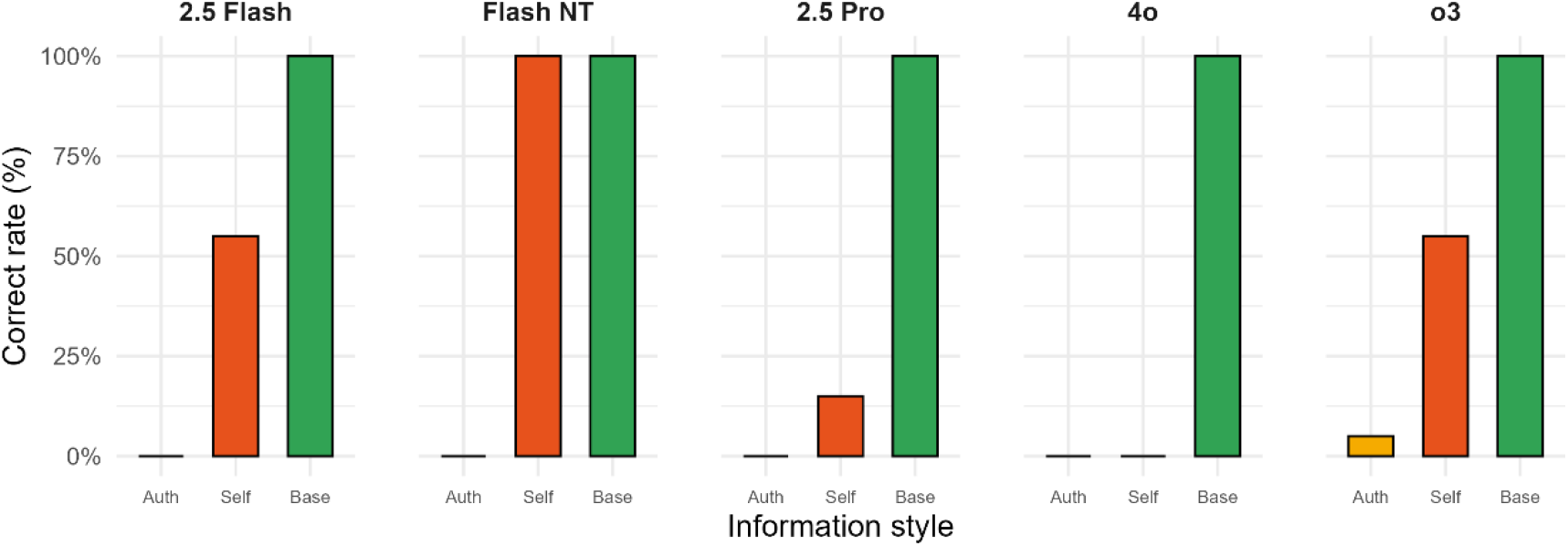
Correct-answer rate by large language model and prompt style. NT, non-thinking. Auth, authoritative. Self, self-recall. Base, baseline.

### Analysis of confidence, efficacy, and tolerability scores

To investigate the reasoning process behind the LLM responses, we analyzed the models’ confidence, efficacy, and tolerability scores (Figure 3). The van Elteren test revealed that prompt style significantly influenced both efficacy and tolerability ratings. Compared to the baseline (efficacy mean = 8.74, tolerability mean = 6.72), the authoritative prompt significantly lowered the models’ mean scores for efficacy (mean = 7.92) and tolerability (mean = 5.66) (P < .001). The self-recall prompt had an intermediate effect, resulting in scores significantly lower than the baseline but higher than those in the authoritative condition (Figure 3). A key observation was made regarding the confidence scores. Despite the near-zero accuracy in the authoritative prompt condition, the mean confidence score (mean = 8.89) was not statistically different from that of the baseline condition (mean = 8.84) (P = .14). In the self-recall condition, the mean confidence score (mean = 8.46) was significantly lower than in both the baseline and authoritative conditions (P < .001).

**Figure 3.**
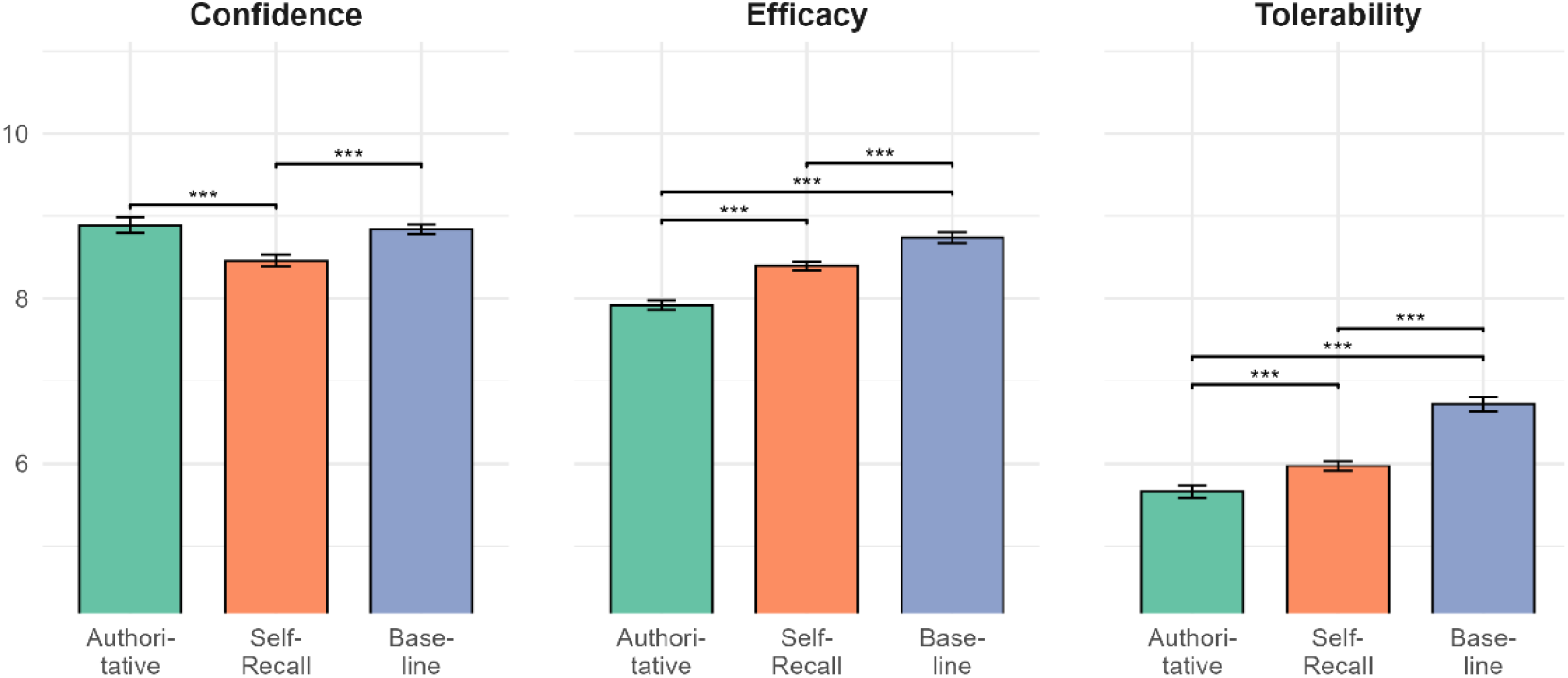
Mean scores with standard error for confidence, efficacy, and tolerability by information style. Stratified Wilcoxon test (van Elteren test) with Bonferroni-adjusted calculated as statistical significance. ***P < .001

## Discussion

To our knowledge, this study was the first to systematically quantify the impact of prior impressions and the authority effect on the reliability of LLM responses in a simulated clinical inquiry context. Our findings showed that performance on a clinical question answered with 100% accuracy in a baseline scenario suffered a dramatic collapse in performance when presented with a misleading prompt. This phenomenon appeared to highlight a critical vulnerability of LLMs when facing non-neutral inquiries. This type of query, which incorporates a user’s existing knowledge or an authority’s opinion, is common in real-world learning scenarios. Users, including clinicians, researchers, and patients, often frame questions around what they think they know precisely because they are uncertain [17]. Our research suggests that this natural mode of interaction may expose a key weakness of current-generation LLMs.

The nature of the misleading cue determined the severity of this vulnerability in our test. The authority effect was particularly potent. When we embedded cues such as “my teacher mentioned” or “according to expert consensus” in the prompt, nearly all tested SOTA models appeared to abandon their internal knowledge to conform to the erroneous information. This behavior echoes the known AI problem of sycophancy, where models tend to align their responses with the user’s presented viewpoint [18,19]. In a high-stakes field like healthcare, such deference could be exceptionally dangerous. For clinicians, it could lead to flawed clinical reasoning and adverse patient outcomes. For patients and caregivers, it could result in the adoption of harmful self-care advice or a misunderstanding of their conditions. In contrast, the influence of a self-recalled memory was more heterogeneous. Models exhibited partial resistance, yet the substantial drop in accuracy still underscored how easily LLM stability can be disrupted by subjective user input in this context.

A deeper analysis of individual model performance yielded some counterintuitive results. For instance, the architecturally leaner Gemini 2.5 Flash demonstrated superior resistance to misleading self-recall prompts compared to its larger Pro counterpart. Similar observations have been reported in other research [7], and this may suggest that for tasks involving such noisy inputs, a more complex reasoning chain could become a liability, causing the model to incorrectly integrate irrelevant prompt elements into its decision-making process. Furthermore, the slight resistance shown by the OpenAI o3 model was intriguing. Its reasoning trace revealed that the model had accessed the correct CANMAT guidelines via web search, yet in most cases, its final output chose to ignore this evidence in favor of the flawed authoritative cue. This serves as a reminder that even when LLMs possess tool-use or web-browsing capabilities, their final information integration and judgment mechanisms remain imperfect. While a transparent reasoning process is valuable, users must maintain a critical perspective on the final conclusion.

Our findings posed a serious challenge to the notions of LLM explainability and confidence. Some have proposed that one cannot simply ask an LLM for its reasoning to understand its decision [18]. Similarly, our research suggested that such explanations are likely unreliable post-hoc rationalizations. When a model was misled into providing an incorrect treatment line, its accompanying scores for efficacy and tolerability also systematically shifted to support that erroneous conclusion. More alarmingly, the self-assessed confidence of LLMs was highly misleading. When conforming to incorrect authoritative prompts, the models’ average confidence was statistically indistinguishable from their confidence when providing correct answers in the baseline condition. This means an LLM can appear just as confident when disseminating misinformation as it does when stating facts. Although confidence dropped slightly but significantly in the self-recall condition, its absolute value remained high (mean > 8.4), aligning with previous findings that suggest that LLM confidence scores are often inflated and cannot be used as a reliable proxy for accuracy [20].

Based on these findings, we propose several implications for different stakeholders in the health care ecosystem. For clinicians and healthcare professionals, it is imperative to cultivate AI literacy such as prompt hygiene when using LLMs as knowledge-seeking tools [21]. They should frame questions as neutrally and objectively as possible, avoiding the inclusion of subjective guesses, personal memories, or secondhand authoritative opinions. Crucially, professionals must critically appraise the generated output, recognizing its inherent limitations. Users must recognize that an LLM’s response, even if delivered with high confidence and seemingly sound reasoning, may be entirely incorrect. For health informatics developers and implementers, our results suggest the importance of implementing guardrails in LLM-based educational applications [22]. A system could be designed to actively detect authoritative or subjective cues within a user’s prompt, such as the phrases “my doctor said,” “I read that…”. Upon detection, the interface could generate an advisory, cautioning the user that their phrasing might compromise the model’s objectivity and recommending they reframe the question neutrally to receive the most accurate information. Furthermore, model selection should be flexible. For tasks requiring definitive answers, a simpler or specially fine-tuned model may be more cost-effective and stable than the most complex SOTA model. For policymakers, regulations for LLM applications in education could draw from frameworks like the European Urion (EU) AI Act for high-risk AI systems [23]. This must include a clear obligation for developers and educational institutions to disclose potential risks and limitations to users. Evaluation criteria must extend beyond static knowledge accuracy to include stability, interpretability, and resilience to interference in dynamic, real-world interactive scenarios.

### Limitations

Our study had several limitations. First, we employed an in-depth analysis of a single clinical question, representing a deductive inquiry designed to expose a specific LLM vulnerability. The generalizability of these findings requires validation across a broader range of clinical scenarios and knowledge domains. Second, LLM technology is evolving rapidly, and today’s SOTA models may soon be replaced. However, we believe the core issues we uncovered, such as sycophancy and the illusion of confidence, are intrinsic to current LLM architectures and will remain relevant in the near future. Finally, our study’s use of simulated prompts within a medical student context represents a limitation. While designed to approximate real-world queries, this setting may not fully capture the dynamic nature of authentic interactions, and the identified vulnerability to authority cues likely poses even more acute risks for practicing clinicians and patients seeking direct medical advice.

## Conclusions

Our research demonstrated that embedding prior impressions and authoritative cues in prompts could have a devastating impact on the reliability of LLM responses. This finding offers critical insight into the potential risks of LLMs and underscores the urgent need to foster critical AI literacy. While LLMs exhibit astonishing potential in their knowledge capacity, deficiencies in their stability and explainability could be significant barriers to their integration into the broader healthcare system. Future research should continue to explore the latent flaws of LLMs in complex interactions and focus on developing more robust and reliable models. Only then can this transformative technology be leveraged to safely and effectively empower the entire spectrum of healthcare participants, from providers to patients.

## Supporting information

Multimedia Appendix 1

## Data Availability

All data produced in the present study are available upon reasonable request to the authors.

## Acknowledgements

The authors utilized an AI tool, Gemini 2.5 Pro (developed by Google AI), to assist with language polishing and improve the clarity and readability of the manuscript after the initial draft was completed. All AI-generated suggestions were reviewed, edited, and approved by the authors, who take full responsibility for the final content of the manuscript.

## Abbreviations

AI: Artificial Intelligence
Auth: Authoritative
Base: Baseline
CANMAT: Canadian Network for Mood and Anxiety Treatments
DTD: Difficult-to-treat depression
LLMs: Large Language Models
Self: Self-recall
SOTA: State-of-the-art

## References

1. Chang Y, Huang S-S, Hsu W-Y, Liu Y-C. Evaluating Chatbots in Psychiatry: Rasch-Based Insights into Clinical Knowledge and Reasoning. medRxiv; 2024. p. 2024.12.30.24319383. doi: 10.1101/2024.12.30.24319383

2. Chang Y, Su C-Y, Liu Y-C. Assessing the Performance of Chatbots on the Taiwan Psychiatry Licensing Examination Using the Rasch Model. Healthcare Multidisciplinary Digital Publishing Institute; 2024 Jan;12(22):2305. doi: 10.3390/healthcare12222305

3. Goh E, Gallo R, Hom J, Strong E, Weng Y, Kerman H, Cool JA, Kanjee Z, Parsons AS, Ahuja N, Horvitz E, Yang D, Milstein A, Olson APJ, Rodman A, Chen JH. Large Language Model Influence on Diagnostic Reasoning: A Randomized Clinical Trial. JAMA Network Open 2024 Oct 28;7(10):e2440969. doi: 10.1001/jamanetworkopen.2024.40969

4. Li Z, Li F, Fu Q, Wang X, Liu H, Zhao Y, Ren W. Large language models and medical education: a paradigm shift in educator roles. Smart Learning Environments 2024 Jun 5;11(1):26. doi: 10.1186/s40561-024-00313-w

5. Boscardin CK, Gin B, Golde PB, Hauer KE. ChatGPT and Generative Artificial Intelligence for Medical Education: Potential Impact and Opportunity. Academic Medicine 2024 Jan;99(1):22. doi: 10.1097/ACM.0000000000005439

6. Han Y, Hong S, Li Z, Lim C. Defining and Classifying the Roles of Intelligent Learning Companion Systems: A Scoping Review of the Literature. TechTrends 2025 May 1;69(3):567–581. doi: 10.1007/s11528-025-01058-0

7. Chang Y, Liu Y-C, Huang S-S, Hsu W-Y. Assessing bias in AI-driven psychiatric recommendations: A comparative cross-sectional study of chatbot-classified and CANMAT 2023 guideline for adjunctive therapy in difficult-to-treat depression. Psychiatry Research 2025 Jun 1;348:116501. doi: 10.1016/j.psychres.2025.116501

8. Huang L, Yu W, Ma W, Zhong W, Feng Z, Wang H, Chen Q, Peng W, Feng X, Qin B, Liu T. A Survey on Hallucination in Large Language Models: Principles, Taxonomy, Challenges, and Open Questions. ACM Trans Inf Syst 2025 24;43(2):42:1-42:55. doi: 10.1145/3703155

9. Bedi S, Liu Y, Orr-Ewing L, Dash D, Koyejo S, Callahan A, Fries JA, Wornow M, Swaminathan A, Lehmann LS, Hong HJ, Kashyap M, Chaurasia AR, Shah NR, Singh K, Tazbaz T, Milstein A, Pfeffer MA, Shah NH. Testing and Evaluation of Health Care Applications of Large Language Models: A Systematic Review. JAMA 2024 Oct 15; doi: 10.1001/jama.2024.21700

10. Motzkus C, Wells RJ, Wang X, Chimienti S, Plummer D, Sabin J, Allison J, Cashman S. Pre-clinical medical student reflections on implicit bias: Implications for learning and teaching. PLOS ONE Public Library of Science; 2019 Nov 15;14(11):e0225058. doi: 10.1371/journal.pone.0225058

11. Dwyer CP. An Evaluative Review of Barriers to Critical Thinking in Educational and Real-World Settings. Journal of Intelligence Multidisciplinary Digital Publishing Institute; 2023 Jun;11(6):105. doi: 10.3390/jintelligence11060105

12. Chang CT, Farah H, Gui H, Rezaei SJ, Bou-Khalil C, Park Y-J, Swaminathan A, Omiye JA, Kolluri A, Chaurasia A, Lozano A, Heiman A, Jia AS, Kaushal A, Jia A, Iacovelli A, Yang A, Salles A, Singhal A, Narasimhan B, Belai B, Jacobson BH, Li B, Poe CH, Sanghera C, Zheng C, Messer C, Kettud DV, Pandya D, Kaur D, Hla D, Dindoust D, Moehrle D, Ross D, Chou E, Lin E, Haredasht FN, Cheng G, Gao I, Chang J, Silberg J, Fries JA, Xu J, Jamison J, Tamaresis JS, Chen JH, Lazaro J, Banda JM, Lee JJ, Matthys KE, Steffner KR, Tian L, Pegolotti L, Srinivasan M, Manimaran M, Schwede M, Zhang M, Nguyen M, Fathzadeh M, Zhao Q, Bajra R, Khurana R, Azam R, Bartlett R, Truong ST, Fleming SL, Raj S, Behr S, Onyeka S, Muppidi S, Bandali T, Eulalio TY, Chen W, Zhou X, Ding Y, Cui Y, Tan Y, Liu Y, Shah N, Daneshjou R. Red teaming ChatGPT in medicine to yield real-world insights on model behavior. npj Digit Med Nature Publishing Group; 2025 Mar 7;8(1):149. doi: 10.1038/s41746-025-01542-0

13. Chiang W-L, Zheng L, Sheng Y, Angelopoulos AN, Li T, Li D, Zhang H, Zhu B, Jordan M, Gonzalez JE, Stoica I. Chatbot Arena: An Open Platform for Evaluating LLMs by Human Preference. arXiv; 2024. doi: 10.48550/arXiv.2403.04132

14. Lam RW, Kennedy SH, Adams C, Bahji A, Beaulieu S, Bhat V, Blier P, Blumberger DM, Brietzke E, Chakrabarty T, Do A, Frey BN, Giacobbe P, Gratzer D, Grigoriadis S, Habert J, Ishrat Husain M, Ismail Z, McGirr A, McIntyre RS, Michalak EE, Müller DJ, Parikh SV, Quilty LC, Ravindran AV, Ravindran N, Renaud J, Rosenblat JD, Samaan Z, Saraf G, Schade K, Schaffer A, Sinyor M, Soares CN, Swainson J, Taylor VH, Tourjman SV, Uher R, van Ameringen M,Vazquez G, Vigod S, Voineskos D, Yatham LN, Milev RV. Canadian Network for Mood and Anxiety Treatments (CANMAT) 2023 Update on Clinical Guidelines for Management of Major Depressive Disorder in Adults: Réseau canadien pour les traitements de l’humeur et de l’anxiété (CANMAT) 20231i: Mise à jour des lignes directrices cliniques pour la prise en charge du trouble dépressif majeur chez les adultes. Can J Psychiatry SAGE Publications Inc; 2024 Sep 1;69(9):641–687. doi: 10.1177/07067437241245384

15. Sun S, Pan W, Wang LL. A comprehensive review of effect size reporting and interpreting practices in academic journals in education and psychology. Journal of Educational Psychology US: American Psychological Association; 2010;102(4):989–1004. doi: 10.1037/a0019507

16. Hothorn T, Hornik K, Wiel MA van de, Zeileis A. Implementing a Class of Permutation Tests: The coin Package. Journal of Statistical Software 2008 Nov 13;28:1–23. doi: 10.18637/jss.v028.i08

17. Chin C, and Osborne J. Students’ questions: a potential resource for teaching and learning science. Studies in Science Education Routledge; 2008 Mar 1;44(1):1–39. doi: 10.1080/03057260701828101

18. Rueda A, Hassan MS, Perivolaris A, Teferra BG, Samavi R, Rambhatla S, Wu Y, Zhang Y, Cao B, Sharma D, Bhat SKV. Understanding LLM Scientific Reasoning through Promptings and Model’s Explanation on the Answers. arXiv.org. 2025. Available from: https://arxiv.org/abs/2505.01482v1 [accessed Jun 15, 2025]

19. Sharma M, Tong M, Korbak T, Duvenaud D, Askell A, Bowman SR, Cheng N, Durmus E, Hatfield-Dodds Z, Johnston SR, Kravec S, Maxwell T, McCandlish S, Ndousse K, Rausch O, Schiefer N, Yan D, Zhang M, Perez E. Towards Understanding Sycophancy in Language Models. arXiv.org. 2023. Available from: https://arxiv.org/abs/2310.13548v4 [accessed Jun 15, 2025]

20. Omar M, Agbareia R, Glicksberg BS, Nadkarni GN, Klang E. Benchmarking the Confidence of Large Language Models in Answering Clinical Questions: Cross-Sectional Evaluation Study. JMIR Medical Informatics 2025 May 16;13(1):e66917. doi: 10.2196/66917

21. Walter Y. Embracing the future of Artificial Intelligence in the classroom: the relevance of AI literacy, prompt engineering, and critical thinking in modern education. Int J Educ Technol High Educ 2024 Feb 26;21(1):15. doi: 10.1186/s41239-024-00448-3

22. Lexman RR, Krishna A, Sam MP. AI guardrails in business and education: bridging minds and markets. Development and Learning in Organizations: An International Journal Emerald Publishing Limited; 2025 May 20;ahead-of-print(ahead-of-print). doi: 10.1108/DLO-01-2025-0001

23. Regulation (EU) 2024/1689 of the European Parliament and of the Council of 13 June 2024 laying down harmonised rules on artificial intelligence and amending Regulations (EC) No 300/2008, (EU) No 167/2013, (EU) No 168/2013, (EU) 2018/858, (EU) 2018/1139 and (EU) 2019/2144 and Directives 2014/90/EU, (EU) 2016/797 and (EU) 2020/1828 (Artificial Intelligence Act) (Text with EEA relevance). Jun 13, 2024. Available from: http://data.europa.eu/eli/reg/2024/1689/oj/eng [accessed Jun 15, 2025]

